# Safety, usability, patient comfort, and exploratory biofilm outcomes of the superhydrophilic antifouling LubriShield® Foley catheter: a randomised controlled trial

**DOI:** 10.64898/2026.09.29.26364294

**Authors:** Ana I. Romero, Soumya Madhusudhan, Mathangi Krishnakumar, Ramya Sambasivam, Serhiy Surkov, Per Wirsén, Linda Bergström, Graeme Brookes, Karin Leire, Jan Andersson

## Abstract

**Background:** Indwelling urinary catheters can cause pain, urethral irritation and trauma and provide surfaces for microbial colonisation and biofilm formation. The LubriShield® Foley catheter incorporates a covalently bonded, superhydrophilic antifouling hydrogel designed to reduce surface friction and limit biofilm accumulation without releasing an antimicrobial agent. This study evaluated its safety and clinical performance compared with an uncoated silicone Foley catheter, including usability and patient comfort, and explored catheter-associated microbial colonisation and biofilm formation.

**Methods:** This prospective, randomised, single-blind, single-centre controlled clinical investigation enrolled 33 adults requiring indwelling urinary catheterisation for 72 hours to 14 days. Participants received either the LubriShield® Foley catheter (n = 16) or an uncoated Sterimed all-silicone Foley catheter (n = 17). The primary endpoint was safety based on adverse-event reporting; the prespecified safety criterion was the absence of a serious adverse device effect in the LubriShield® group. Secondary endpoints included investigator-rated ease and functionality of catheter insertion and removal and participant-reported pain and irritation/discomfort assessed using 0–10 numerical rating scales. Urine microbiology, catheter colonisation and catheter-level biofilm measurements were exploratory endpoints. Analyses were performed in the as-treated population. Numerical rating scale outcomes were compared using two-sided Mann–Whitney U tests; exploratory biofilm analyses used permutation tests with bootstrap confidence intervals.

**Results:** No serious adverse events or serious adverse device effects occurred. Nine non-serious adverse events were reported and resolved completely. At least one adverse event occurred in 4/16 (25.0%) participants in the LubriShield® group and 4/17 (23.5%) participants in the control group. The prespecified safety criterion, defined as the absence of a SADE in the LubriShield® group, was met. LubriShield® was rated more favourably for ease of insertion (*p* = 0.0096), functionality during insertion (*p* = 0.0051), ease of removal (*p* = 0.0037) and functionality during removal (*p* = 0.0085). Participants reported significantly less pain and irritation/discomfort during insertion, while the catheter was indwelling and during removal (all *p* < 0.05). Outcomes 7 ± 1 days after catheter removal were similar between groups. Absolute biofilm biomass and bacterial and fungal loads did not differ significantly. In a post hoc exploratory normalised analysis, the biofilm-to-bacterial-load ratio was lower with LubriShield® (median 0.052 vs 0.080; permutation *p* = 0.043), although the bootstrap 95% confidence interval for the between-group difference included zero.

**Conclusions:** The LubriShield® Foley catheter demonstrated a favourable short-term safety profile and was associated with improved investigator-rated usability and participant-reported comfort compared with an uncoated silicone Foley catheter. A post hoc exploratory analysis was compatible with reduced biofilm accumulation relative to bacterial load, but this finding is hypothesis-generating only and requires confirmation in larger, adequately powered studies with prespecified biofilm endpoints.

**Trial registration:** Clinical Trials Registry–India (CTRI), CTRI/2025/05/086528, prospectively registered on 7 May 2025, before participant enrolment. Subsequently registered with ClinicalTrials.gov, NCT07070921.

## Background

Urinary catheterisation is essential for bladder drainage in hospitalised patients but is associated with complications including pain, urethral trauma, catheter-associated urinary tract infection (CAUTI), encrustation, and biofilm formation (1–3). An estimated 15–25% of hospitalised patients undergo urinary catheterisation during their hospital stay, and approximately 75% of hospital-acquired urinary tract infections are catheter-associated (4). Reducing urinary catheter-related morbidity while maintaining effective urinary drainage therefore remains an important clinical objective.

Microbial colonisation and biofilm formation contribute substantially to catheter-associated complications. Following attachment to the catheter surface, microorganisms can develop structured biofilms embedded within an extracellular polymeric matrix that promotes persistence and tolerance to host defences and antimicrobial treatment (5–7). Biofilm development can contribute to persistent colonisation, infection, encrustation, catheter obstruction and device failure (8–11). Strategies to limit these processes have included antibiotic-impregnated catheters, silver-alloy coatings, antiseptic agents and other antimicrobial surface modifications (12,13). However, improvements in clinically relevant outcomes have been inconsistent, and antimicrobial approaches may exert selective pressure favouring antimicrobial resistance (14).

Non-antimicrobial antifouling surfaces offer an alternative strategy by modifying interactions between biological material, microorganisms, and the catheter surface rather than directly killing microorganisms (9,15). Hydrophilic and superhydrophilic coatings maintain a highly hydrated interfacial layer that can reduce friction and nonspecific protein adsorption and may interfere with stable microbial attachment and subsequent biofilm development (16–18). These properties could potentially address both mechanical catheter-related discomfort and microbial fouling without releasing an antimicrobial agent.

The LubriShield® Foley catheter is an all-silicone catheter with a covalently bonded superhydrophilic antifouling hydrogel coating on its internal and external surfaces. The coating is designed to provide persistent surface hydration, reduce friction and limit biofilm development without antimicrobial release (19). In preclinical studies, LubriShield® reduced surface friction and biofilm formation by common uropathogens under static and dynamic conditions, including in the presence of the host-derived protein fibrinogen, without substantially reducing the number of viable surface-associated microorganisms (19). These findings are consistent with an antifouling rather than directly antimicrobial mechanism. A previous randomised pilot study in 30 participants undergoing transurethral resection of bladder tumour found the catheter to be well tolerated during short-term use, with no device-related adverse events and no significant differences in microbiological outcomes compared with a standard silicone Foley catheter (20).

Although the pilot study provided initial evidence of clinical safety and feasibility during short-term catheterisation, the clinical performance of LubriShield® during longer indwelling use had not been established. In particular, further evaluation was needed to assess device safety, usability, and patient comfort over a broader catheterisation period and to explore whether the antifouling properties observed preclinically were detectable on catheters following clinical use.

The present prospective randomised controlled trial therefore evaluated the safety and clinical performance of the LubriShield® Foley catheter compared with an uncoated silicone Foley catheter in adults requiring indwelling urinary catheterisation for 72 hours to 14 days. The primary objective was to evaluate device safety. Secondary objectives were to assess investigator-rated catheter usability and functionality and participant-reported pain and irritation/discomfort. Urinary microbial colonisation, catheter-associated microbial burden, and biofilm formation were evaluated as exploratory outcomes.

## Materials and Methods

### Study design

This prospective, randomised, single-blind, single-centre, parallel-group controlled clinical investigation compared the safety and clinical performance of the LubriShield® Foley catheter with an uncoated silicone Foley catheter. Participants were blinded to treatment allocation. Healthcare professionals performing catheter insertion and removal were not blinded, and investigator-reported usability assessments were therefore unblinded. Laboratory personnel performing biofilm and microbiological analyses were blinded to treatment allocation.

The study was conducted at St. John’s Medical College Hospital, Bengaluru, India, from June to November 2025 in accordance with ISO 14155:2020, Good Clinical Practice and the principles of the Declaration of Helsinki. The study protocol was approved by the Institutional Ethics Committee of St. John’s Medical College Hospital (IEC reference CT-07/2024), and all participants provided written informed consent before enrolment.

The primary objective was to evaluate device safety. Secondary objectives were to assess catheter usability and functionality and participant-reported pain and irritation/discomfort. Urinary microbial colonisation, catheter-associated microbial burden, and biofilm formation were prespecified exploratory outcomes.

The study was prospectively registered with the Clinical Trials Registry–India (CTRI; CTRI/2025/05/086528) on 7 May 2025, before enrolment of the first participant and was subsequently registered with ClinicalTrials.gov, NCT07070921.

### Study population

Adults aged 18-84 years who required indwelling urinary catheterisation for at least 72 hours and up to 14 days and were suitable for catheterisation with a 14-, 16- or 18-Fr Foley catheter were eligible. Participants were required to be conscious at enrolment and able to provide written informed consent.

Exclusion criteria were pregnancy or breastfeeding; critical illness defined by an Acute Physiology and Chronic Health Evaluation II (APACHE II) score ≥28; known allergy or sensitivity to silicone; previous participation in the study; concurrent participation in another clinical investigation considered likely to influence the primary endpoint; urinary catheterisation for more than 48 hours before enrolment; symptomatic genitourinary pathology; or anticipated inability to comply with the study protocol, as judged by the principal investigator.

### Randomisation and sample size

Participants were allocated in a 1:1 ratio to receive either a LubriShield® Foley catheter or an uncoated Sterimed all-silicone Foley catheter using a computer-generated centralised randomisation procedure.

The Clinical Investigation Plan specified a planned sample size of 30 participants, with 15 participants per group, for this initial evaluation of device safety, usability and clinical performance. No formal hypothesis-based sample-size calculation was performed, and the study was not powered to demonstrate efficacy, detect uncommon adverse events (AEs) or establish between-group differences in microbiological or biofilm outcomes. A total of 33 participants were ultimately enrolled and randomised, with 16 allocated to the LubriShield® group and 17 to the Sterimed group.

### Investigational and control devices

The investigational device was the LubriShield® Foley catheter, an all-silicone Foley catheter with a proprietary, covalently bonded superhydrophilic antifouling hydrogel coating applied to the internal and external catheter surfaces. The coating is permanent and non-releasing and is intended to reduce surface friction and limit microbial fouling and biofilm development without releasing an antimicrobial agent.

The control device was a commercially available uncoated Sterimed all-silicone Foley catheter. Both devices were used for transurethral bladder drainage in accordance with their intended use.

### Catheterisation procedure

Urinary catheterisation was performed by trained healthcare professionals using standard aseptic transurethral insertion procedures. After insertion into the bladder, the retention balloon was inflated with sterile water and the catheter was connected to a urinary drainage bag.

Catheters remained in situ according to clinical need for a minimum of 72 hours and a maximum of 14 days. Participants were followed for 7 ± 1 days after catheter removal.

### Safety assessments

Safety was evaluated based on the occurrence, severity, seriousness and assessed relationship to the study device or catheterisation procedure of AEs, serious adverse events (SAEs) and serious adverse device effects (SADEs). AEs were recorded throughout the catheterisation period and through the 7 ± 1-day post-removal follow-up. In accordance with the prespecified Statistical Analysis Plan, the safety hypothesis was considered met if no SADEs were observed in the LubriShield® group.

### Usability, functionality, and participant-reported assessments

Catheter usability and functionality, and participant-reported pain and irritation/discomfort were assessed using 11-point numerical rating scales ranging from 0 to 10, with lower scores indicating more favourable outcomes.

Investigators assessed ease of catheter insertion, catheter functionality during insertion, ease of understanding and following the instructions for use, ease of catheter removal and catheter functionality during removal.

Participants assessed pain and irritation/discomfort during insertion, while the catheter was in situ and during removal. At the 7 ± 1-day follow-up, participants also assessed pain, irritation/discomfort and urinary retention.

### Urinary microbiological assessment

Urine dipstick testing and urine sampling were performed before catheterisation and before catheter removal. Urine samples were cultured quantitatively to assess urinary microbial colonisation. Bacteriuria was defined, in accordance with the Clinical Investigation Plan, as growth of one or more bacterial species at ≥10⁵ colony-forming units (CFU)/mL (≥10⁸ CFU/L) in urine culture. Bacterial and fungal isolates were identified using standard microbiological procedures.

### Catheter biofilm and microbiological analysis

Following catheter removal, six 2-cm segments were collected from three predefined catheter regions. Segments A and B were obtained near the catheter tip, with segment A positioned below the eyelet; segments C and D were obtained from the middle region; and segments E and F were obtained near the catheter hub. Within each region, adjacent paired segments were assigned to complementary biofilm and microbiological analyses (Fig. 1). Segments A, C and E were used for biofilm biomass quantification, whereas segments B, D and F were used for viable microbial enumeration.

**Figure 1.**
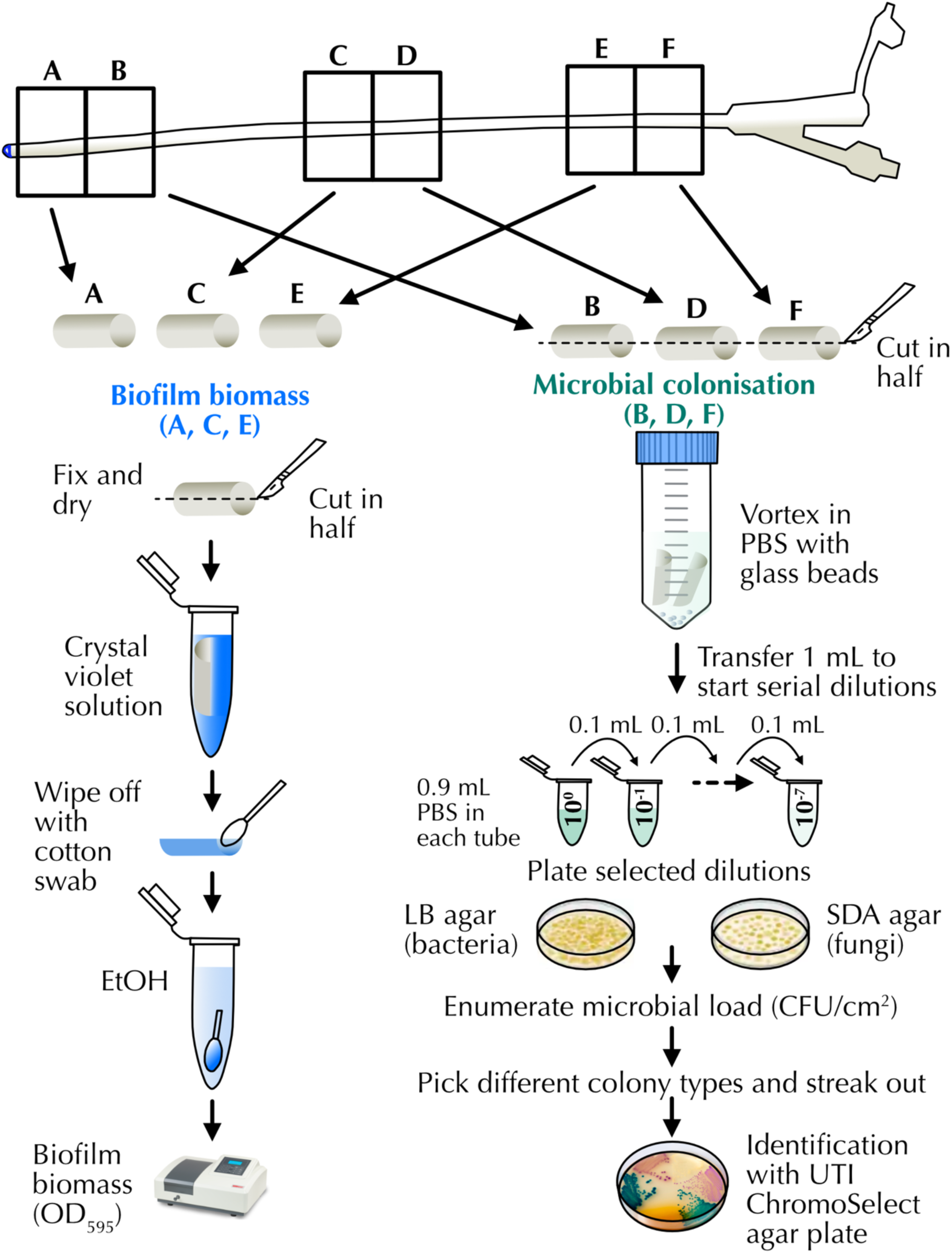
Catheter sampling and analytical workflow for biofilm biomass and microbial burden. Following catheter removal, three predefined catheter regions were sampled as adjacent paired segments (A/B, C/D and E/F). Segments A, C and E were used to quantify surface-associated biofilm biomass by crystal violet staining and measurement of optical density at 595 nm (OD₅₉₅). Adjacent segments B, D and F were used to quantify recoverable viable microorganisms. Surface-associated microorganisms were recovered by vortexing catheter segments in phosphate-buffered saline (PBS) with glass beads, followed by serial dilution and culture on Luria–Bertani (LB) agar for bacterial enumeration and Sabouraud dextrose agar (SDA) for fungal enumeration. Microbial burden was expressed as colony-forming units per square centimetre (CFU/cm²) of catheter surface. Representative colonies displaying distinct morphologies were further evaluated on UTI ChromoSelect agar for presumptive bacterial identification.

For biofilm biomass analysis, segments A, C and E were sequentially fixed in 1% glutaraldehyde in phosphate-buffered saline (PBS), 60% ethanol and 80% ethanol and then dried. Each segment was cut longitudinally, and one half was stained with 0.04% crystal violet. Following rinsing and drying, stained material was recovered from the inner catheter surface using a PBS-moistened cotton swab and extracted in 96% ethanol. Biofilm biomass was quantified spectrophotometrically by measuring optical density at 595 nm (OD₅₉₅).

For microbiological analysis, segments B, D and F were cut longitudinally and vortexed in PBS containing glass beads to recover surface-associated microorganisms. Serial dilutions were plated on Luria–Bertani agar for bacterial enumeration and Sabouraud dextrose agar for fungal enumeration and incubated at 37°C for 24–48 hours. Viable microbial burden was expressed as colony-forming units per square centimetre of catheter surface (CFU/cm²). Representative colonies of different morphologies were further evaluated using UTI ChromoSelect agar for presumptive bacterial identification.

For catheter-level analyses, measurements obtained from the three predefined catheter regions were combined to derive one representative value per catheter for biofilm biomass and bacterial and fungal burden.

A post hoc exploratory analysis examined biofilm biomass relative to recoverable microbial burden by normalising catheter-level OD₅₉₅ values to the corresponding bacterial or fungal burden. Samples with no detectable microbial growth were retained in the microbial-load analyses but excluded from the corresponding ratio analysis because a ratio could not be calculated.

### Outcome measures

The primary endpoint was device safety, assessed from the incidence and characteristics of AEs in accordance with ISO 14155:2020.

Secondary endpoints were investigator-rated catheter usability and functionality during insertion and removal and participant-reported pain and irritation/discomfort during insertion, while the catheter was in situ, during removal and at follow-up.

Prespecified exploratory endpoints included culture-confirmed bacteriuria, defined as ≥10⁵ CFU/mL in urine culture, catheter-associated bacterial and fungal burden and catheter-associated biofilm biomass.

A post hoc exploratory endpoint evaluated catheter-level biofilm biomass relative to bacterial and fungal burden.

### Statistical analysis

Data were analysed using Microsoft Excel 2021 and R statistical software (version 4.4.1). Participant characteristics, device exposure, safety outcomes, investigator-rated usability and functionality, participant-reported outcomes, urinary microbiological findings, and catheter-associated biofilm and microbiological measurements were summarised descriptively. Continuous variables are presented as means with standard deviations (SDs) or medians with interquartile ranges (IQRs), as appropriate, and categorical variables as numbers and percentages. Analyses were conducted in the as-treated population, defined as all enrolled participants who received a study catheter. Missing values were not imputed, and the number of observations contributing to each analysis was reported where applicable.

Given the small sample size and ordinal nature of the numerical rating scale (NRS) outcomes, between-group comparisons were performed using two-sided Mann–Whitney U tests. Effect sizes were estimated using the Vargha–Delaney A statistic with 95% confidence intervals. A was defined as the probability that a randomly selected LubriShield® participant had a higher NRS score than a randomly selected Sterimed participant, with half-weight assigned to ties. As lower NRS scores indicated more favourable outcomes, A < 0.5 favoured LubriShield®, A > 0.5 favoured Sterimed, and A = 0.5 indicated no between-group tendency. Confidence intervals for A were calculated using a non-parametric variance estimate derived from the pairwise comparison scores and a two-sided normal-approximation, with confidence limits restricted to the interval [0,1].

Biofilm and microbiological outcomes were analysed at the catheter level, with each catheter contributing one independent observation. For each catheter, OD₅₉₅ measurements from the three predefined biofilm segments were averaged to derive a catheter-level measure of biofilm biomass. Corresponding CFU/cm² measurements were log₁₀-transformed, and measurements from the three predefined microbiological segments were averaged to derive catheter-level bacterial and fungal burdens. Between-group differences in these catheter-level outcomes were estimated as differences in group medians. Two-sided permutation tests based on the between-group difference in medians were used for between-group comparisons, with 95% confidence intervals for the median differences estimated by bootstrap resampling.

In a post hoc exploratory analysis, catheter-level biofilm biomass was evaluated relative to the corresponding recoverable bacterial or fungal burden, expressed as the ratio of mean OD₅₉₅ to mean log₁₀ CFU/cm². Between-group differences in these normalised measures were evaluated using the same permutation and bootstrap-resampling procedures. Samples with no detectable microbial growth were retained as zero in microbial-burden analyses but excluded from the corresponding ratio analysis because the ratio was not computable.

All statistical tests were two-sided, and no adjustment was made for multiple comparisons. Microbiological and biofilm outcomes were prespecified as exploratory, whereas analyses of biofilm biomass relative to microbial burden were post hoc. Given the small sample size, the exploratory analyses were interpreted considering effect estimates, 95% confidence intervals, and the direction and magnitude of observed differences, in addition to *p* values. Exact *p* values are reported where applicable.

## Results

### Study population and baseline characteristics

Between 2 June and 21 November 2025, 33 participants were enrolled and randomised: 16 to the LubriShield® Foley catheter group and 17 to the Sterimed uncoated silicone Foley catheter group. All participants received the allocated catheter and were included in the as-treated analysis. Participant flow through the study is shown in the CONSORT flow diagram (Fig. 2).

**Figure 2.**
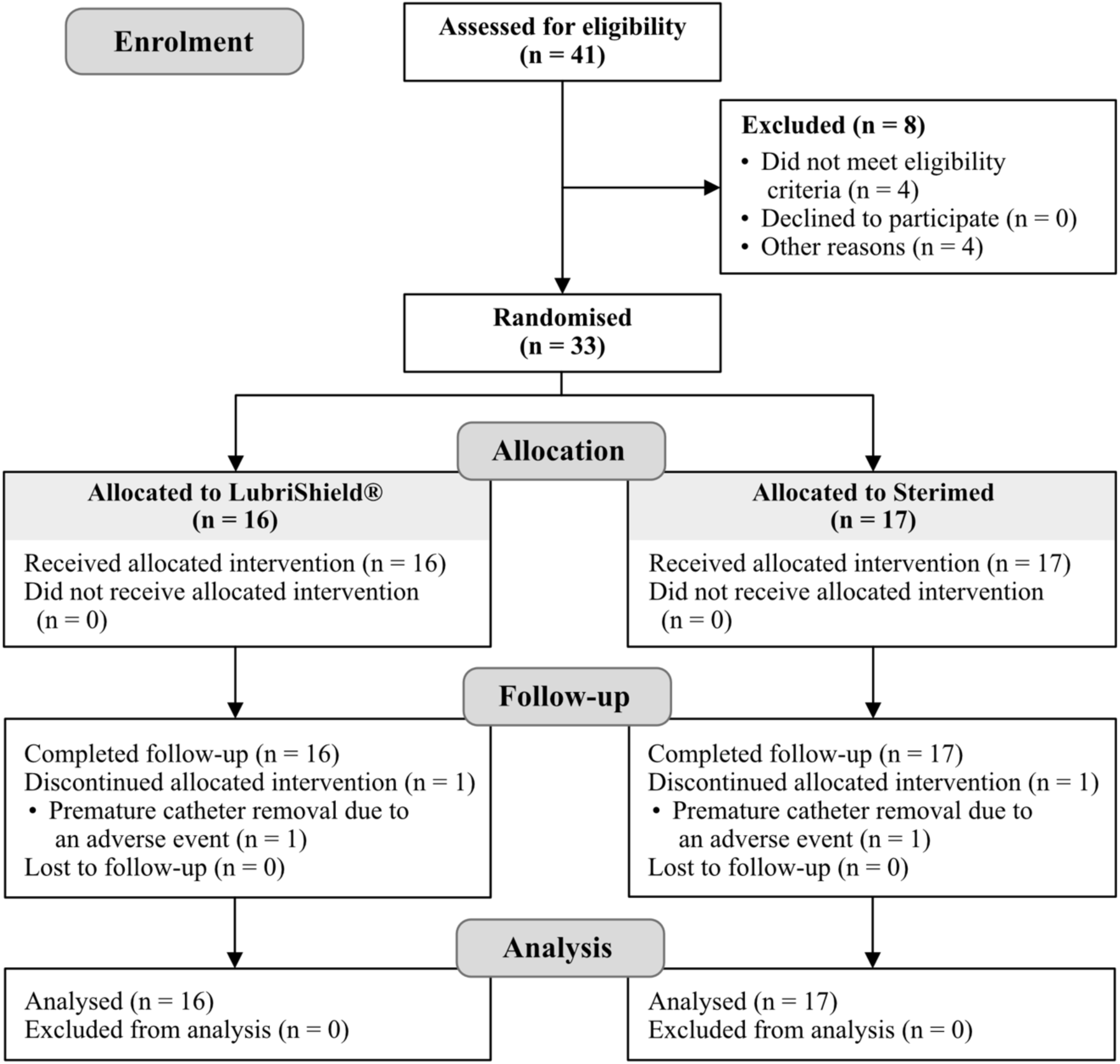
CONSORT flow diagram of participant enrolment, allocation, follow-up and analysis. Thirty-three participants were randomised, with 16 allocated to the LubriShield® group and 17 to the Sterimed group. All participants received their allocated intervention and were included in the analysis. One participant in each group discontinued the allocated intervention prematurely because of an adverse event but completed study follow-up. No participants were lost to follow-up.

Baseline demographic and clinical characteristics are shown in table 1. Overall, 20/33 participants (60.6%) were male and 13/33 (39.4%) were female. Nine of 16 participants (56.3%) in the LubriShield® group and 11/17 (64.7%) in the Sterimed group were male. The most common age category was 31–50 years, comprising 6/16 (37.5%) participants in the LubriShield® group and 7/17 (41.2%) in the Sterimed group.

**Table 1.**
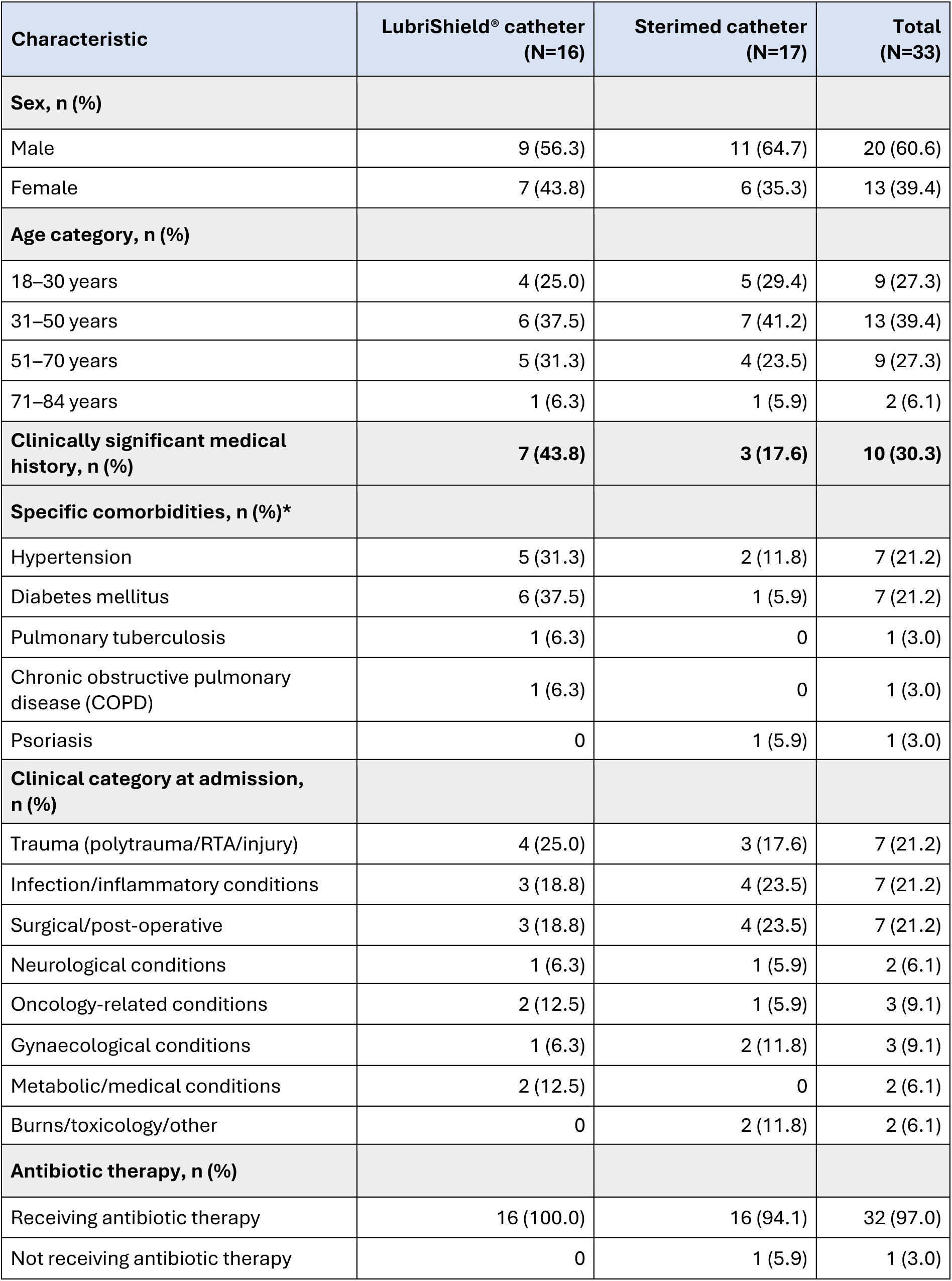
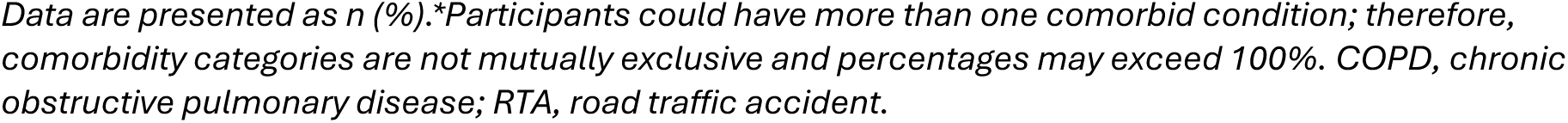
Baseline demographic and clinical characteristics by treatment group.

| Characteristic | LubriShield® catheter<br>(N=16) | Sterimed catheter<br>(N=17) | Total<br>(N=33) |
| --- | --- | --- | --- |
| **Sex, n (%)** |  |  |  |
| Male | 9 (56.3) | 11 (64.7) | 20 (60.6) |
| Female | 7 (43.8) | 6 (35.3) | 13 (39.4) |
| **Age category, n (%)** |  |  |  |
| 18–30 years | 4 (25.0) | 5 (29.4) | 9 (27.3) |
| 31–50 years | 6 (37.5) | 7 (41.2) | 13 (39.4) |
| 51–70 years | 5 (31.3) | 4 (23.5) | 9 (27.3) |
| 71–84 years | 1 (6.3) | 1 (5.9) | 2 (6.1) |
| **Clinically significant medical history, n (%)** | **7 (43.8)** | **3 (17.6)** | **10 (30.3)** |
| **Specific comorbidities, n (%)\*** |  |  |  |
| Hypertension | 5 (31.3) | 2 (11.8) | 7 (21.2) |
| Diabetes mellitus | 6 (37.5) | 1 (5.9) | 7 (21.2) |
| Pulmonary tuberculosis | 1 (6.3) | 0 | 1 (3.0) |
| Chronic obstructive pulmonary disease (COPD) | 1 (6.3) | 0 | 1 (3.0) |
| Psoriasis | 0 | 1 (5.9) | 1 (3.0) |
| **Clinical category at admission, n (%)** |  |  |  |
| Trauma (polytrauma/RTA/injury) | 4 (25.0) | 3 (17.6) | 7 (21.2) |
| Infection/inflammatory conditions | 3 (18.8) | 4 (23.5) | 7 (21.2) |
| Surgical/post-operative | 3 (18.8) | 4 (23.5) | 7 (21.2) |
| Neurological conditions | 1 (6.3) | 1 (5.9) | 2 (6.1) |
| Oncology-related conditions | 2 (12.5) | 1 (5.9) | 3 (9.1) |
| Gynaecological conditions | 1 (6.3) | 2 (11.8) | 3 (9.1) |
| Metabolic/medical conditions | 2 (12.5) | 0 | 2 (6.1) |
| Burns/toxicology/other | 0 | 2 (11.8) | 2 (6.1) |
| **Antibiotic therapy, n (%)** |  |  |  |
| Receiving antibiotic therapy | 16 (100.0) | 16 (94.1) | 32 (97.0) |
| Not receiving antibiotic therapy | 0 | 1 (5.9) | 1 (3.0) |

*Data are presented as n (%). \*Participants could have more than one comorbid condition; therefore, comorbidity categories are not mutually exclusive and percentages may exceed 100%. COPD, chronic obstructive pulmonary disease; RTA, road traffic accident.*

The study population was clinically heterogeneous, reflecting the range of conditions requiring intensive care and urinary catheterisation. Clinically significant medical history was reported in 7/16 participants (43.8%) in the LubriShield® group and 3/17 (17.6%) in the Sterimed group. Hypertension was reported in 5/16 (31.3%) and 2/17 (11.8%) participants, respectively, and diabetes mellitus in 6/16 (37.5%) and 1/17 (5.9%), respectively. Concomitant antibiotic therapy was administered to 16/16 participants (100%) in the LubriShield® group and 16/17 (94.1%) in the Sterimed group.

### Device exposure

A 14-Fr catheter was used in all 16 participants (100%) in the LubriShield® group and 16/17 (94.1%) in the Sterimed group; one participant (5.9%) in the Sterimed group received an 18-Fr catheter. Mean catheterisation duration was 5.19 (SD 2.04) days in the LubriShield® group and 5.41 (SD 2.60) days in the Sterimed group; median duration was 5 days in both groups. Catheterisation lasted 3–6 days in 12/16 participants (75.0%) and 12/17 (70.6%), respectively. One participant in each group underwent catheter removal before 72 hours because of an adverse event.

### Urine microbiology

No bacterial growth was detected in pre-catheterisation urine cultures from any of the 33 participants. Positive baseline urine dipstick findings were recorded in 5/16 participants (31.3%) in the LubriShield® group and 5/17 (29.4%) in the Sterimed group.

At catheter removal, urine culture results were available for 15/16 participants (93.8%) in the LubriShield® group and 16/17 (94.1%) in the Sterimed group. Culture-confirmed bacteriuria (≥10⁵ CFU/mL) was detected in 3/15 participants (20.0%) in the LubriShield® group and 2/16 (12.5%) in the Sterimed group; cultures were negative in 12/15 (80.0%) and 14/16 (87.5%), respectively. All five episodes of bacteriuria were recorded as AEs.

### Safety outcomes

Nine AEs were reported during the study: five in the LubriShield® group and four in the Sterimed group. These occurred in 4/16 (25.0%) participants in the LubriShield® group and 4/17 (23.5%) participants in the Sterimed group. All AEs were non-serious and resolved completely without sequelae. No SAEs or SADEs were reported. The prespecified safety criterion, defined as the absence of a SADE in the LubriShield® group, was met.

In the LubriShield® group, three participants experienced bacteriuria, assessed as possibly device-related and probably procedure-related. Two episodes required no intervention, whereas one was managed by modification of antibiotic therapy. One additional participant experienced pain and haematuria following a technically difficult catheter insertion involving multiple attempts. Both events were assessed as probably device-related and possibly procedure-related. The pain was classified as severe and led to catheter removal and analgesic treatment.

In the Sterimed group, two participants experienced bacteriuria, assessed as possibly device-related. One episode required no intervention, whereas the other was managed by modification of antibiotic therapy. One participant experienced device-related discomfort that led to precautionary catheter removal. Fever occurred in one participant and was assessed as unrelated to the device or catheterisation procedure.

No new AEs or delayed complications were reported during the 7 ± 1-day post-removal follow-up.

### Usability and participant-reported outcomes

#### Catheter insertion

Ease of understanding and following the instructions for use during catheter insertion was rated similarly for the two devices, with a median score of 0 in both groups (A = 0.570, 95% CI 0.435– 0.705; *p* = 0.320).

Investigator-reported ease of catheter insertion scores were lower with LubriShield® than with Sterimed (median 1 vs 2; A = 0.241, 95% CI 0.065–0.417; *p* = 0.0096; Fig. 3A), as were catheter functionality scores during insertion (median 1 vs 2; A = 0.222, 95% CI 0.060–0.384; *p* = 0.0051; Fig. 3B).

**Figure 3.**
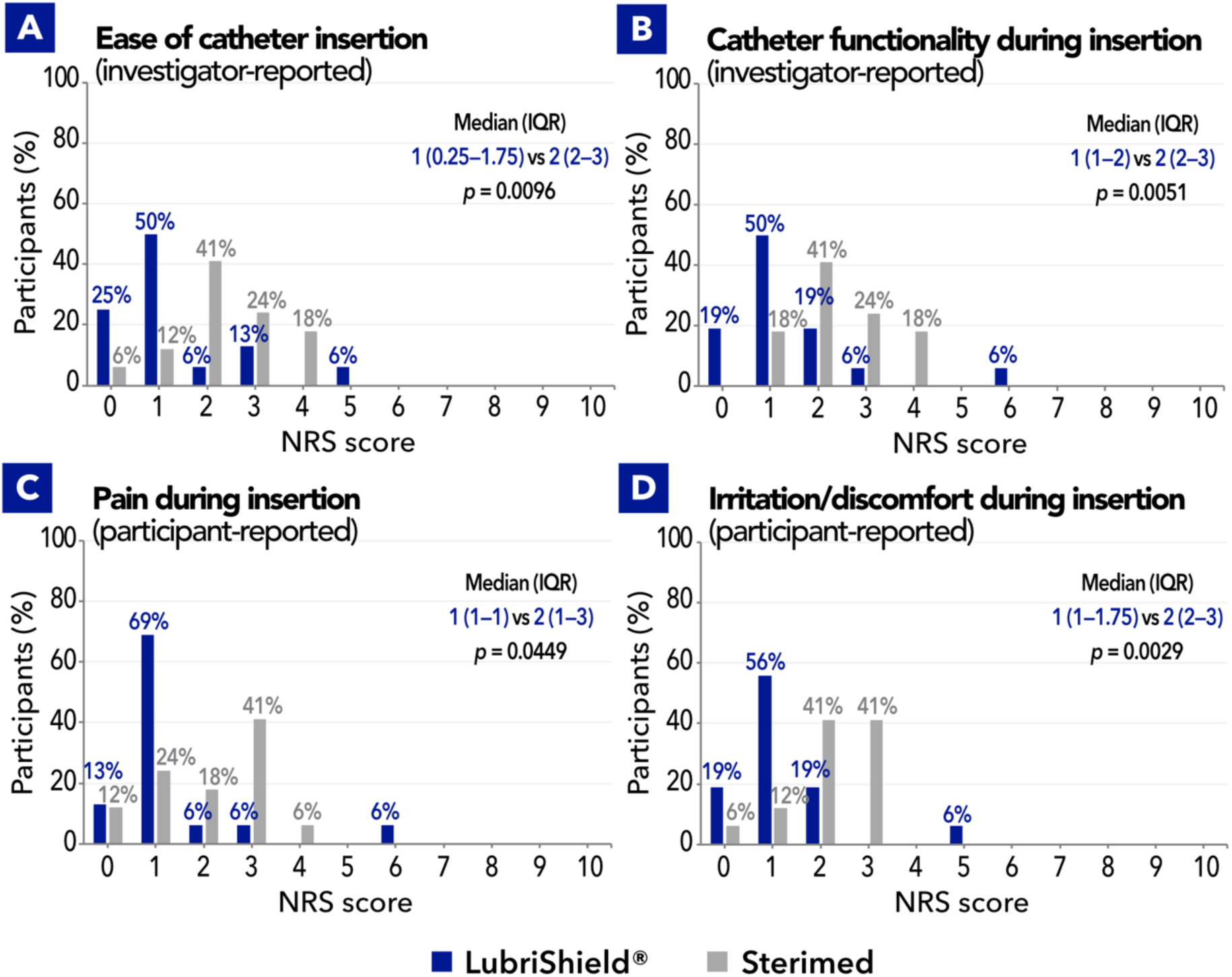
Investigator- and participant-reported assessments during catheter insertion. Distributions of numerical rating scale (NRS) scores for (A) investigator-reported ease of catheter insertion, (B) investigator-reported catheter functionality during insertion, (C) participant-reported pain during insertion and (D) participant-reported irritation/discomfort during insertion. Bars represent the percentage of participants at each NRS score, with percentages shown above the bars. Lower scores indicate more favourable assessments (0=most favourable; 10=least favourable). Median (IQR) values and two-sided p values from Mann–Whitney U tests are shown for each assessment. Vargha–Delaney A effect sizes with 95% confidence intervals are reported in the text; A < 0.5 favours LubriShield® because lower NRS scores indicate more favourable assessments.

Participants reported lower scores with LubriShield® for pain during insertion (median 1 vs 2; A = 0.305, 95% CI 0.120–0.490; *p* = 0.0449; Fig. 3C) and irritation/discomfort during insertion (median 1 vs 2; A = 0.206, 95% CI 0.043–0.368; *p* = 0.0029; Fig. 3D).

#### Indwelling period and catheter removal

While the catheter was in situ, participants reported lower scores with LubriShield® than with Sterimed for pain (median 1 vs 2; A = 0.217, 95% CI 0.046–0.387; *p* = 0.0051; Fig. 4A) and irritation/discomfort (median 0.5 vs 2.5; A = 0.162, 95% CI 0.019–0.305; *p* < 0.001; Fig. 4B).

**Figure 4.**
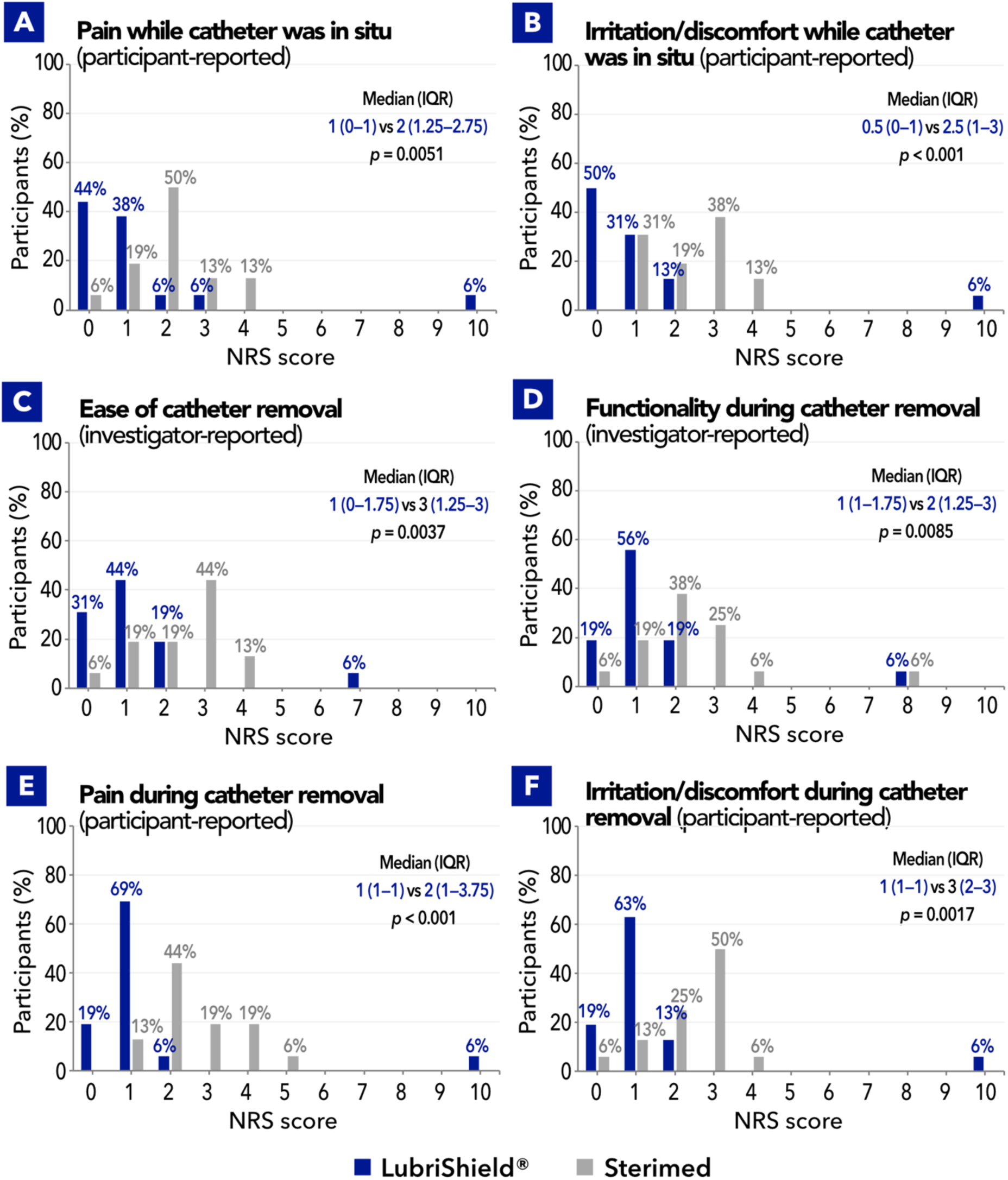
Investigator- and participant-reported assessments during catheterisation and catheter removal. Distributions of numerical rating scale (NRS) scores for (A) participant-reported pain while the catheter was in situ, (B) participant-reported irritation/discomfort while the catheter was in situ, (C) investigator-reported ease of catheter removal, (D) investigator-reported catheter functionality during removal, (E) participant-reported pain during catheter removal and (F) participant-reported irritation/discomfort during catheter removal. Bars represent the percentage of participants at each NRS score, with percentages shown above the bars. Lower scores indicate more favourable assessments (0=most favourable; 10=least favourable). Median (IQR) values and two-sided p values from Mann–Whitney U tests are shown for each assessment. Vargha–Delaney A effect sizes with 95% confidence intervals are reported in the text; A < 0.5 favours LubriShield® because lower NRS scores indicate more favourable assessments.

During catheter removal, ease of understanding and following the instructions for use was rated similarly between groups, with a median score of 0 in both groups (A = 0.594, 95% CI 0.450– 0.738; *p* = 0.216). Investigator-reported ease of catheter removal scores were lower with LubriShield® than with Sterimed (median 1 vs 3; A = 0.205, 95% CI 0.042–0.368; *p* = 0.0037; Fig. 4C), as were functionality scores during removal (median 1 vs 2; A = 0.236, 95% CI 0.067–0.405; *p* = 0.0085; Fig. 4D).

Participants also reported lower scores with LubriShield® for pain during catheter removal (median 1 vs 2; A = 0.127, 95% CI 0.000–0.264; *p* < 0.001; Fig. 4E) and irritation/discomfort during removal (median 1 vs 3; A = 0.186, 95% CI 0.022–0.349; *p* = 0.0017; Fig. 4F).

#### Follow-up after catheter removal

At the 7 ± 1-day follow-up, participant-reported pain, irritation/discomfort and urinary retention scores were low in both groups, with no statistically significant between-group differences. Median pain scores were 1 in both groups (A = 0.581, 95% CI 0.396–0.766; *p* = 0.3876; Fig. 5A), as were median irritation/discomfort scores (A = 0.544, 95% CI 0.359–0.729; *p* = 0.6502; Fig. 5B) and urinary retention scores (A = 0.548, 95% CI 0.355–0.741; *p* = 0.6233; Fig. 5C).

**Figure 5.**
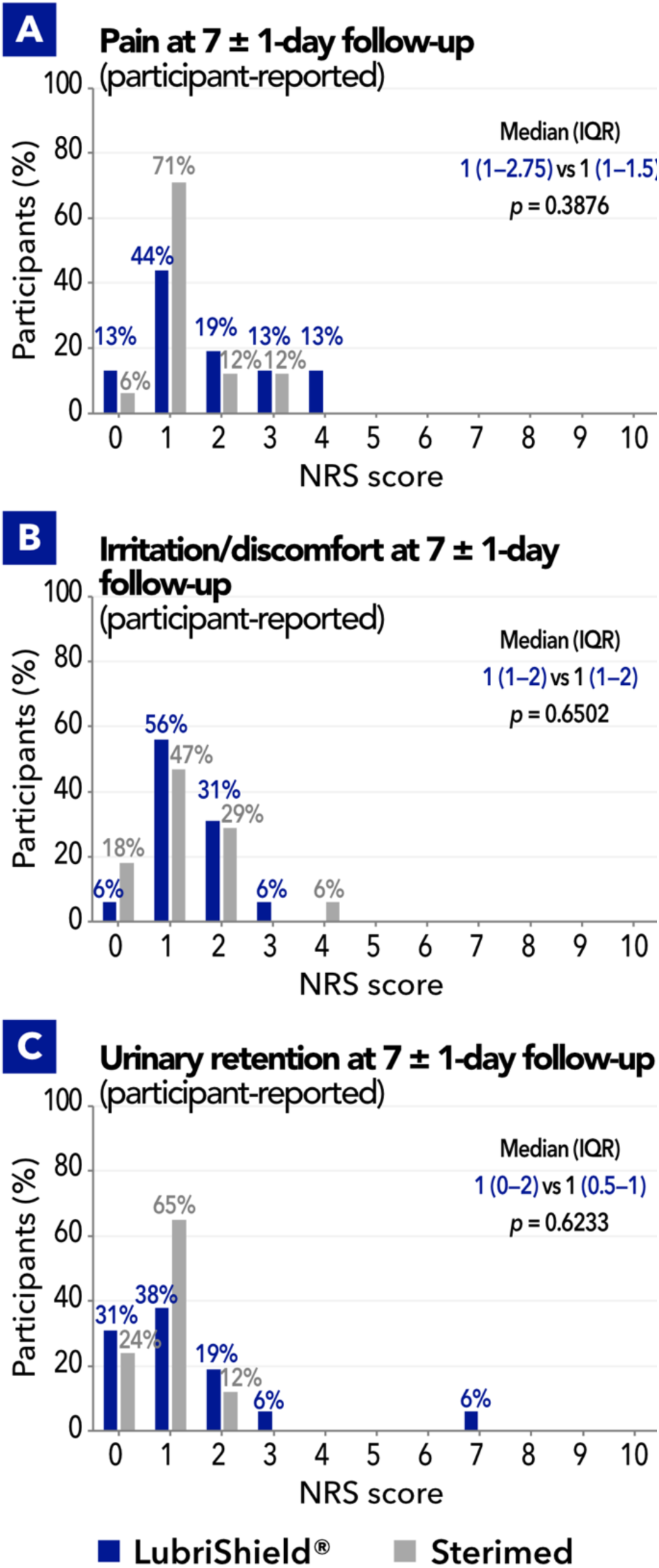
Participant-reported outcomes at the 7 ± 1-day follow-up after catheter removal. Distributions of numerical rating scale (NRS) scores for (A) pain, (B) irritation/discomfort and (C) urinary retention at the 7 ± 1-day post-removal follow-up. Bars represent the percentage of participants at each NRS score, with percentages shown above the bars. Lower scores indicate more favourable outcomes (0=most favourable; 10=least favourable). Median (IQR) values and two-sided p values from Mann–Whitney U tests are shown for each assessment. Vargha–Delaney A effect sizes with 95% confidence intervals are reported in the text; A < 0.5 favours LubriShield® because lower NRS scores indicate more favourable assessments.

### Catheter-surface biofilm and microbial burden

#### Biofilm biomass

Catheter-level biofilm biomass was lower in the LubriShield® group than in the Sterimed group (median OD₅₉₅, 0.249 vs 0.312; Fig. 6A). The estimated between-group median difference was −0.063 (bootstrap 95% CI −0.211 to 0.046) with no statistically significant difference between groups (permutation *p* = 0.253).

**Figure 6.**
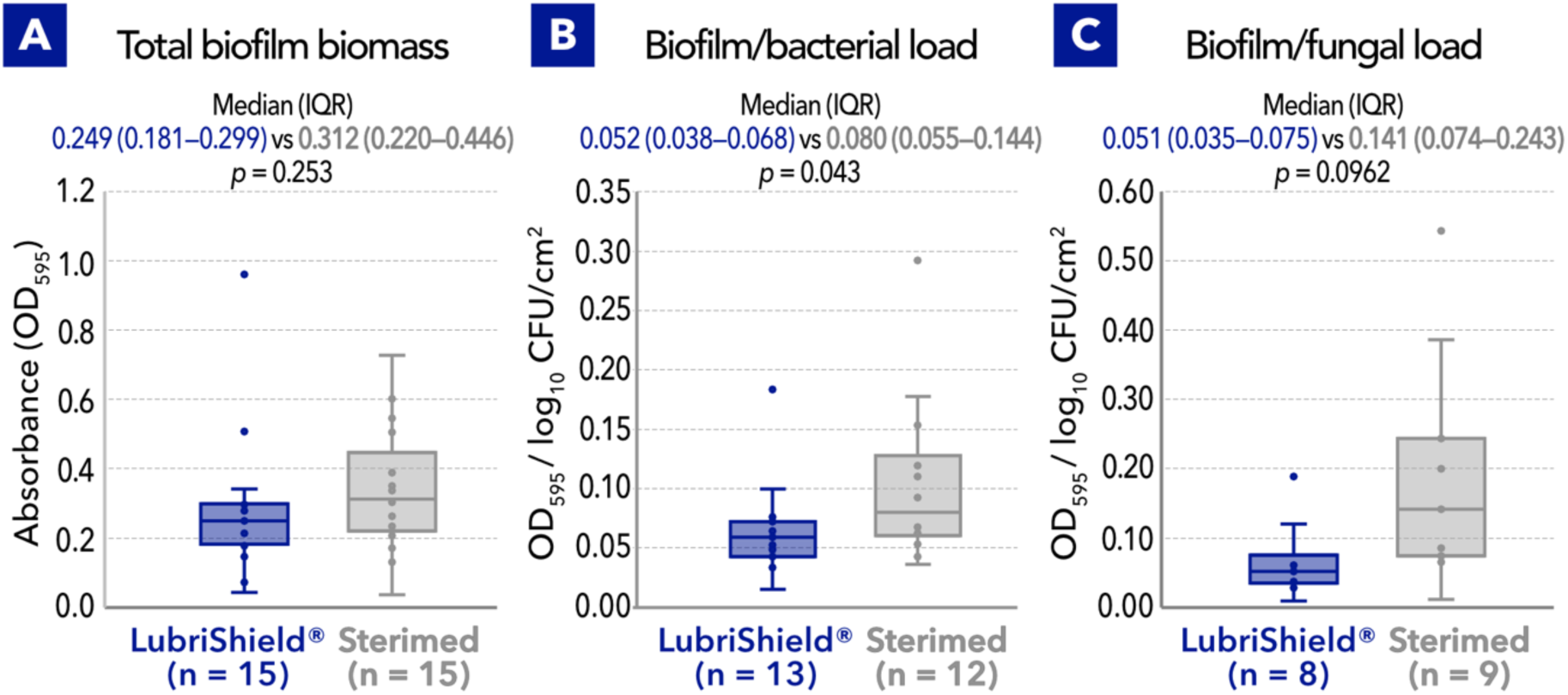
Catheter-surface biofilm biomass and biofilm biomass relative to microbial burden. (A) Catheter-level biofilm biomass measured by crystal violet staining and expressed as OD₅₉₅. (B) Biofilm biomass relative to bacterial burden recovered on Luria–Bertani (LB) agar, expressed as OD₅₉₅/log₁₀ CFU/cm². (C) Biofilm biomass relative to fungal burden recovered on Sabouraud dextrose agar (SDA), expressed as OD₅₉₅/log₁₀ CFU/cm². Boxplots show medians and interquartile ranges (IQRs), with individual points representing catheter-level observations. Sample sizes are shown below each group. Values above each panel show median (IQR) values and two-sided p values from permutation tests based on between-group differences in medians. The biofilm-to-bacterial-burden analysis was post hoc and exploratory.

##### Microbial burden

Median bacterial burden recovered from catheter surfaces was 4.905 log₁₀ CFU/cm² with LubriShield® and 4.191 log₁₀ CFU/cm² with Sterimed *(p* = 0.124). Median fungal burden was 2.557 and 1.437 log₁₀ CFU/cm², respectively (*p* = 0.534). Neither bacterial nor fungal burden differed significantly between groups.

Microorganisms recovered from the catheter surfaces included *Enterococcus* spp., *Pseudomonas aeruginosa*, *Klebsiella pneumoniae*, *Staphylococcus aureus*, *Escherichia coli*, and *Candida* spp. The types of microorganisms identified were broadly comparable between groups.

##### Biofilm biomass relative to microbial load

In the post hoc exploratory analysis, biofilm biomass relative to recoverable bacterial burden, expressed as the ratio of OD₅₉₅/log₁₀ CFU/cm², was lower for LubriShield® than for Sterimed catheters (median ratio, 0.052 vs. 0.080; permutation *p* = 0.043; Fig. 6B). The estimated between-group median difference was −0.028 (bootstrap 95% CI −0.0938 to 0.0048). Thus, LubriShield® catheters accumulated less biofilm biomass relative to the bacterial load recovered from the catheter surface, although the bootstrap 95% confidence interval for the between-group median difference included zero.

The biofilm-to-fungal-burden ratio showed the same direction of difference (median 0.051 with LubriShield® vs 0.141 with Sterimed) although the between-group comparison was not statistically significant (permutation *p* = 0.0962; Fig. 6C).

## Discussion

In this randomised controlled trial, the LubriShield® Foley catheter showed a favourable short-term safety profile and was associated with more favourable investigator-reported usability ratings and lower participant-reported pain and irritation/discomfort scores than an uncoated silicone Foley catheter. No SAEs or SADEs occurred, and all reported AEs resolved completely. Differences in usability and participant-reported outcomes were observed during catheter insertion, while the catheter was in situ and during removal, whereas scores at the 7 ± 1-day post-removal follow-up were low in both groups. Exploratory catheter analyses did not provide evidence of lower absolute biofilm biomass or microbial burden with LubriShield®. A post hoc analysis showed a lower biofilm-to-bacterial-burden ratio with LubriShield®, although uncertainty around the effect estimate warrants cautious interpretation.

The usability and comfort findings are consistent with the superhydrophilic and lubricious properties of the permanently bound hydrogel coating. Hydrophilic surfaces maintain a strongly hydrated interfacial layer that can reduce friction between the catheter and urethral mucosa and thereby potentially limit mechanical irritation during insertion and removal (21,22). Similarly, Zhu et al reported lower urethral pain and bladder irritation together with reduced urinary epithelial-cell and leukocyte counts, in participants receiving super-lubricious Foley catheters compared with conventional catheters (23). In the present study, lower pain and irritation/discomfort scores were observed not only during insertion and removal but also while the catheter was in situ. This pattern is compatible with reduced surface friction being relevant throughout catheter exposure, including during catheter movement within the urethra. The absence of an apparent difference at post-removal follow-up is also consistent with an effect associated primarily with the period of catheter exposure.

Safety was the primary objective of this study. Nine AEs were reported overall, occurring in 4/16 (25.0%) participants in the LubriShield® group and 4/17 (23.5%) participants in the Sterimed group. All were non-serious and resolved completely without sequelae, and no SAEs, SADEs, device malfunctions, catheter-associated urinary tract infections or cases of sepsis occurred. These findings provide initial clinical evidence supporting the short-term safety and tolerability of the coated catheter. However, the sample size was insufficient to assess uncommon AEs or establish equivalence in safety between the devices.

The exploratory catheter-surface analyses provide a more nuanced assessment of the proposed antifouling effect. Absolute biofilm biomass was numerically lower in the LubriShield® group (median OD₅₉₅ 0.249 vs 0.312), but the estimated between-group difference was imprecise and its 95% confidence interval included zero. Bacterial and fungal burdens were not lower with LubriShield®. The absence of reduced viable microbial burden is not inconsistent with the intended mechanism of LubriShield®, which is designed as an antifouling rather than antimicrobial surface and is not intended to exert direct microbicidal activity (19).

In the post hoc normalised analysis, the ratio of biofilm biomass to recoverable bacterial burden was lower with LubriShield® than with Sterimed (median 0.052 vs 0.080; permutation *p* = 0.043). The estimated between-group median difference (LubriShield® minus Sterimed) was −0.028; although the bootstrap 95% confidence interval (−0.0938 to 0.0048) included zero. Thus, the permutation test provided evidence of a between-group difference, while the magnitude of this difference was estimated imprecisely and zero remained plausible under the bootstrap procedure. The biofilm-to-fungal-burden ratio showed a difference in the same direction, although this analysis included fewer evaluable catheters and did not show a statistically significant between-group difference. Because both normalised analyses were post hoc, involved relatively small numbers of observations, and were conducted without adjustment for multiple comparisons, these findings should be regarded as hypothesis-generating.

The direction of these exploratory findings is nevertheless biologically compatible with the proposed mechanism of the coating. Biofilm development involves more than the presence of viable microorganisms on a catheter surface; maturation and production of an extracellular polymeric matrix contribute to microbial persistence, antimicrobial tolerance, crystalline deposition, encrustation and obstruction (5–7,24). In our previous in vitro study, LubriShield® reduced biofilm formation by bacterial and fungal uropathogens under static and dynamic conditions, including in the presence of host-derived proteins, without substantially reducing viable surface-associated microorganisms (19). The present findings are therefore consistent with, but do not establish, a mechanism in which the surface interferes with biofilm development rather than directly reducing microbial viability.

Culture-confirmed bacteriuria at catheter removal occurred in 3/15 participants (20.0%) in the LubriShield® group and 2/16 (12.5%) in the Sterimed group. Interpretation is limited by the small number of events, the median catheterisation duration of approximately 5 days and the use of systemic antibiotics in 32/33 participants. The study was not designed or powered to determine an effect on bacteriuria or catheter-associated urinary tract infection, and no conclusions regarding infection prevention can be drawn. Whether modification of catheter-surface biofilm development translates into clinically relevant reductions in infection, encrustation, obstruction or catheter replacement therefore remains to be established.

Several limitations should be considered. First, the small sample size was selected primarily for an initial evaluation of safety and clinical performance and provided limited precision for safety, microbiological and infection-related outcomes. In particular, the absence of SAEs cannot exclude uncommon device-related risks. Second, this was a single-centre study involving a clinically heterogeneous population requiring predominantly short-term catheterisation, which may limit generalisability to other clinical settings and longer-term catheter users. Third, almost all participants received systemic antibiotics, potentially affecting urinary and catheter-surface microbial recovery. Fourth, despite randomisation, clinically significant medical history and some comorbidities were more frequent in the LubriShield® group. Fifth, healthcare professionals were not blinded to device allocation, and investigator-reported usability assessments were therefore susceptible to observer bias, although participants were blinded to treatment allocation. The participant-reported outcomes are therefore less susceptible to this particular source of bias. Finally, the normalised biofilm analyses were post hoc, involved fewer evaluable catheters than the overall study population and were not adjusted for multiple comparisons.

Despite these limitations, the consistency of the participant-reported findings across insertion, the indwelling period, and removal supports further investigation of superhydrophilic surface modification as a means of improving the catheterisation experience. The exploratory catheter-surface findings also provide a hypothesis for future investigation of non-antimicrobial antifouling effects. Larger multicentre randomised trials with longer catheterisation periods, where clinically appropriate, and prespecified biofilm endpoints are needed to confirm these observations and determine whether they translate into clinically important reductions in catheter-associated urinary tract infection, encrustation, obstruction or unplanned catheter replacement.

## Conclusions

The LubriShield® Foley catheter showed a favourable short-term safety profile and was associated with better investigator-rated usability and lower participant-reported pain and irritation/discomfort during catheterisation than an uncoated silicone Foley catheter. No SAEs or SADEs occurred. Exploratory analyses did not establish a reduction in absolute catheter biofilm biomass or microbial burden; however, a post hoc analysis suggested lower biofilm biomass relative to bacterial burden, with uncertainty in the effect estimate. Larger clinical studies are warranted to confirm the observed usability and comfort findings and to determine the clinical relevance of the proposed non-antimicrobial antifouling effect.

## Data Availability

The datasets generated and/or analysed during the current study are not publicly available due to participant confidentiality and data protection requirements but may be available from the corresponding author on reasonable request, subject to applicable ethical, legal and data protection requirements.

## List of abbreviations

AE: adverse event
APACHE II: Acute Physiology and Chronic Health Evaluation II
CAUTI: catheter-associated urinary tract infection
CFU: colony-forming units
CI: confidence interval
COPD: chronic obstructive pulmonary disease
CTRI: Clinical Trials Registry–India
IEC: Institutional Ethics Committee
IQR: interquartile range
LB: Luria–Bertani
NRS: numerical rating scale
OD₅₉₅: optical density at 595 nm
PBS: phosphate-buffered saline
RTA: road traffic accident
SADE: serious adverse device effect
SAE: serious adverse event
SD: standard deviation
SDA: Sabouraud dextrose agar.

## Declarations

### Ethics approval and consent to participate

The study was reviewed and approved by the Institutional Ethics Committee of St. John’s Medical College Hospital, Bengaluru, India (IEC Study Ref. No. CT-07/2024; approval granted 15 November 2024). The study was conducted in accordance with ISO 14155:2020, Good Clinical Practice and the ethical principles of the Declaration of Helsinki. Written informed consent was obtained from all participants before enrolment.

### Consent for publication

Not applicable

### Authors’ contributions

AIR contributed to the biofilm and microbiological analyses, interpretation of the data and findings, and drafting and revision of the manuscript. SM and MK contributed to the conduct of the clinical investigation, acquisition of clinical data, and review of the manuscript. RS contributed to the conduct of the clinical investigation, including participant screening and enrolment, monitoring of patient-related study activities, coordination of study-related laboratory assessments, and review of the manuscript. SS contributed to the biofilm and microbiological analyses, interpretation of the data, and review of the manuscript. PW and LB contributed to the study design, study oversight, interpretation of the findings, and review of the manuscript. GB contributed to the conception and design of the study, study oversight, interpretation of the findings, and revision of the manuscript. KL contributed to the study design, study oversight, and review of the manuscript. JA contributed to the conception and design of the study, interpretation of the findings, and drafting and critical revision of the manuscript. All authors read and approved the final manuscript.

## Acknowledgements

The authors thank the study nurses and medical staff of the Surgical and Neuro ICU, St. John’s Medical College Hospital, Bengaluru, Karnataka, India, for their care of the study participants and assistance with study procedures. The authors also thank Dr. D. Padmanabhan, Meditix Solutions and the Department of Microbiology at St. John’s Medical College Hospital for their contributions to the microbiological and biofilm analyses. The authors gratefully acknowledge Dr. Mohammad Hagsheno for serving as the independent safety officer and Arathy S S, Biostatistician, Dr. Klinisch Research Pvt. Ltd., for performing the statistical analyses. The authors also acknowledge Dr. Klinisch Research Pvt. Ltd., Bengaluru, India, for providing clinical research organisation and study-monitoring services.

